# Lives Saved from Age-Prioritized Covid-19 Vaccination

**DOI:** 10.1101/2021.03.19.21253991

**Authors:** Joshua R. Goldstein, Thomas Cassidy, Ayesha S. Mahmud

## Abstract

**BACKGROUND:** The criteria used to allocate scarce COVID-19 vaccines are hotly contested. While some are pushing just to get vaccines into arms as quickly as possible, others advocate prioritization in terms of risk.

**OBJECTIVE:** Our aim is to use demographic models to show the enormous potential of vaccine risk-prioritization in saving lives.

**METHODS:** We develop a simple mathematical model that accounts for the age distribution of the population and of COVID-19 mortality. This model considers only the direct live-savings for those who receive the vaccine, and does not account for possible indirect effects of vaccination. We apply this model to the United States, Japan, and Bangladesh.

**RESULTS:** In the United States, we find age-prioritization would reduce deaths during a vaccine campaign by about 93 percent relative to no vaccine and 85 percent relative to age-neutral vaccine distribution. In countries with younger age structures, such as Bangladesh, the benefits of age-prioritization are even greater.

**CONTRIBUTION:** For policy makers, our findings give additional support to risk-prioritized allocation of COVID-19 vaccines. For demographers, our results show how the age-structures of the population and of disease mortality combine into an expression of risk concentration that shows the benefits of prioritized allocation. This measure can also be used to study the effects of prioritizing other dimensions of risk such as underlying health conditions.

## Introduction

The availability of highly effective COVID-19 vaccines creates the opportunity for rapidly lowering mortality. A simplified demographic model applied to the United States shows that age-prioritization would lower deaths by 85% relative to an age-neutral vaccine campaign. Even though optimal vaccination is not fully achievable and other considerations need to be taken into account, age-prioritization is an important life-saving benchmark against which alternative vaccination policy can be compared.

The enormous life saving effect of age-prioritization is due to the joint effects of rapidly increasing mortality at older ages (Davies et al., 2020; Goldstein and Lee, 2020), and the relatively small number of elderly (Fig 1 A and B). In this paper, we provide a framework for measuring the benefits of age-prioritization. Our approach considers only the direct effects on saving lives of those who are vaccinated, and does not take into account the effects of reduced viral transmission. This simplified framework can be extended to apply to any other aspect of risk beyond just age.

**Figure 1:**
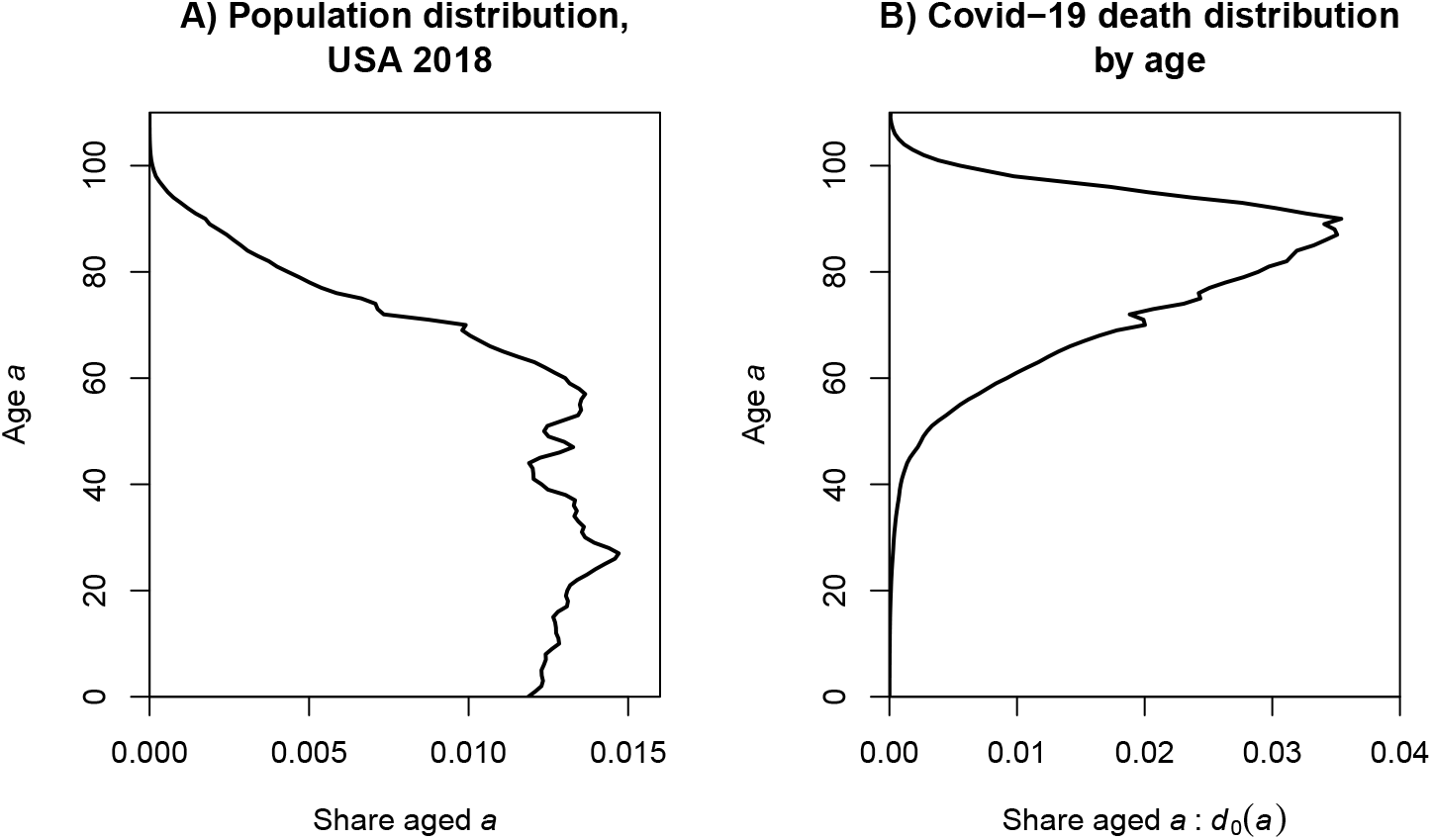
Age structure of the U.S. 2018 population and of Covid-19 deaths.

### A model

To illustrate the life-saving potential of prioritizing vaccines by age, we consider a vaccine campaign that protects a constant share of the population each day until everyone has been vaccinated. We let *p* denote the share of the population that is protected from mortality. With constant shares vaccinated each day, *p* is also equal to fraction of time it takes to vaccinate everyone

We further assume deaths at age *a* would remain at a constant level *d*_0_(*a*) in the absence of vaccination.^1^ During the vaccination campaign, mortality is determined by multiplying the share at each age that remains unvaccinated by the baseline age-specific COVID-19 death rates.

Under an age-neutral campaign, when a share *p* of the population is protected, deaths will be (1 − *p*) of the baseline levels at all ages. Thus,

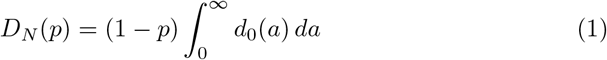

tells us that the ratio of deaths in an age-neutral campaign to baseline, *D*_*N*_ (*p*), will decline linearly from unity to zero over the course of the campaign. The path of the age-neutral campaign is given by the dashed line Figure 2. The ratio of deaths over the entire campaign relative to baseline is the area under this line. This area, denoted *𝒟*_*N*_, corresponds to one-half the deaths that would have occurred without vaccination.

**Figure 2:**
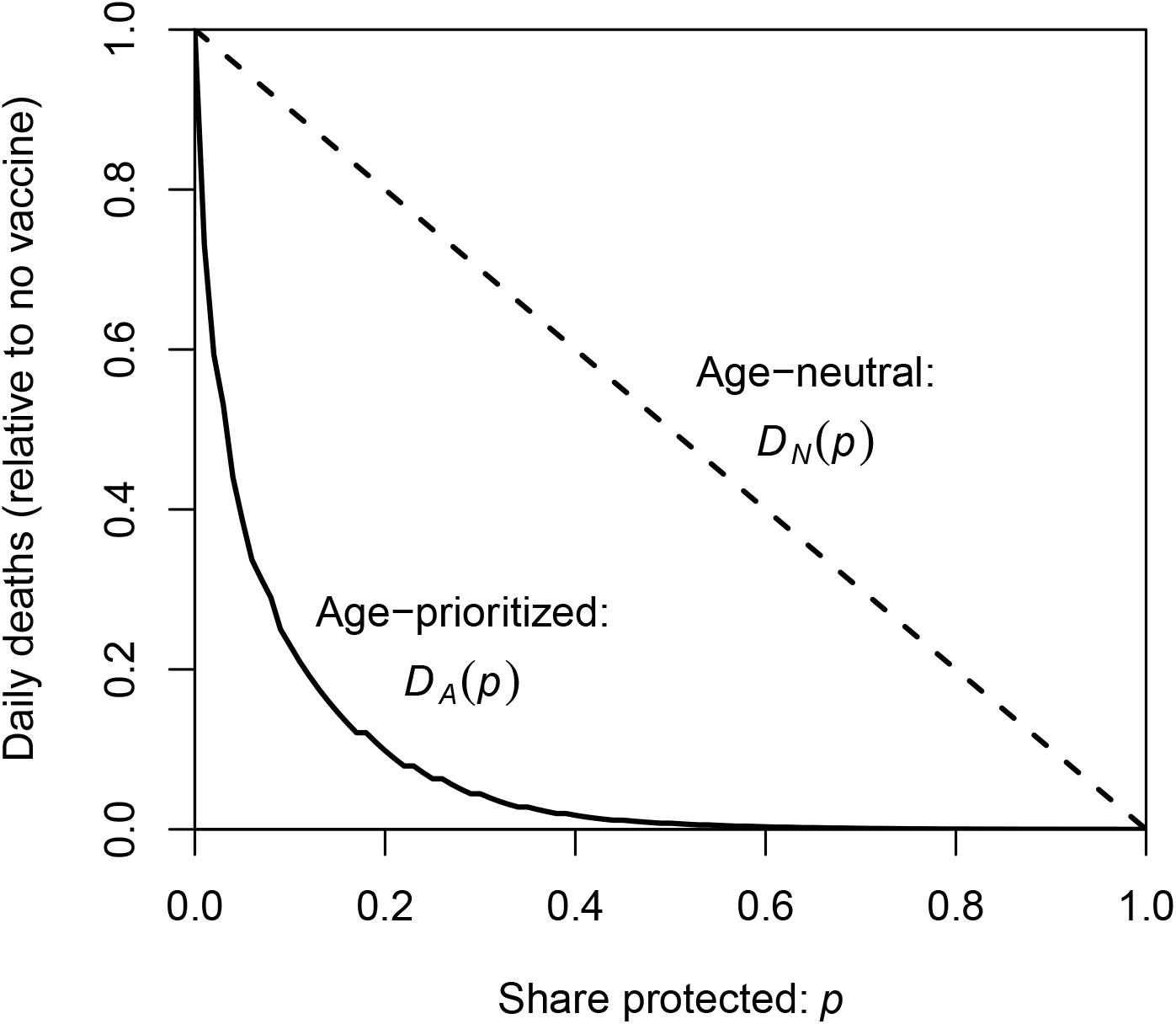
The effectiveness of vaccine prioritization in reducing deaths. The heights of the curves give the deaths, relative to baseline, when a fraction *p* is protected. Under the assumptions of our model, the sum of the area under each curve gives the total deaths over the full course of the campaigns, relative to what would have been observed with no vaccine. The death toll relative to baseline of an age-neutral campaign (*𝒟*_*N*_ = 0.500) shrinks dramatically (*𝒟*_*A*_ = 0.075) under age-prioritization. Source: The *D*_*A*_(*p*) curve is estimated from equation (3) using the U.S. age structure of population and Covid-19 mortality shown in Figure 1.

Now consider an age-prioritized campaign, with deaths declining as the oldest become protected from risk. Let *A*(*p*) be the age over which the share of the population is *p*. In this case, the deaths relative to baseline when a fraction *p* have been protected is

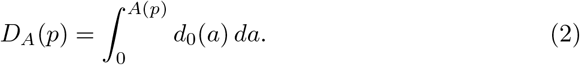

The curve labeled *D*_*A*_(*p*) in Figure 2 shows deaths during an age-prioritized campaign decline quickly at first as the most at-risk are vaccinated and then more slowly thereafter.

Over the entire course of the campaign, the ratio of deaths to baseline can be calculated by summing *D*_*A*_(*p*) over all values of *p*, which increases from 0 to 1 steadily as the campaign goes from start to finish.

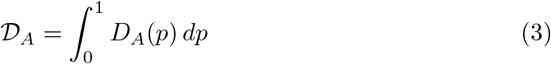

This gives the deaths during an age-prioritized campaign relative to the baseline case without a vaccine. For the United States *𝒟*_*A*_ = 0.075, much smaller than the value of 0.5 of total deaths of a neutral vaccination campaign.

As is shown in the appendix, the effectiveness of age-prioritized vaccination can be re-expressed in terms of *d*_0_(*a*), the baseline age distribution of COVID-19 deaths, and 1 − *F* (*a*), the proportion of the population above age *a*:

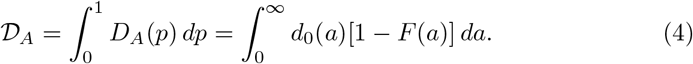

Equation (4) has an intuitive interpretation. The middle term tells us that we can calculate the mortality over the entire time period by summing over the times when a fraction *p* is protected. The right-most term shows that one can also sum over age, with 1 − *F* (*a*) telling us the fraction of the campaign that people aged *a* are exposed, and *d*_0_(*a*) giving the deaths at that age.

## Results

We apply our model to estimates of age-specific COVID-19 death rates based on cumulative reports by CDC and population estimates from U.S. Census (Goldstein and Lee, 2020). A baseline schedule of daily deaths *d*_0_(*a*) at age *a* is obtained by applying these rates to the age-structured population (Fig 1 B).

In the United States, we calculate that *𝒟*_*A*_ is about 0.075. This means that an idealized age-prioritized campaign would lower the lower the death toll by about 92.5% relative to no vaccine, and about 85% relative to the age-neutral campaign. The life-saving effects of age-prioritization can be seen in the much smaller area under the prioritized curve in Fig 2).

To make the magnitudes more concrete, consider a hypothetical campaign lasting 150 days that correspond very roughly to the United States case. Assume that deaths would remain at 2,000 per day over this time in the absence of vaccination. Over these 150 days, 300,000 people would die in the absence of a vaccine, 150,000 would die with age-neutral vaccination, and about 22,000 would die under age-prioritized vaccination.

As vaccination distribution expands in developing countries with younger age-structures, age-prioritization may prove even more effective. The age-structure of COVID-19 mortality risk is not thought to vary enormously across countries, but population pyramids do vary. Applying the same standard age-structure of COVID-19 mortality rates to the observed age-structures, we find that *𝒟*_*A*_*/𝒟*_*N*_ is 0.06 in Bangladesh, 0.26 in Japan (compared to 0.15 in the United States).

Much of the benefit of age-prioritization can be obtained by following strict-prioritization for the oldest segment of the population. For example, strict age-ordering for the first 20 percent of the population with age-neutral open eligibility thereafter is described by tracing along the age-prioritized *D*_*A*_(*p*) curve from *p* = 0 to *p* = 0.2 for the age-prioritized period and then drawing a straight-line (a “chord”) from *D*_*A*_(*p* = .2) to *D*_*A*_(*p* = 1). The area under partially age-prioritized curve for the United States is about 0.100, as compared to 0.075 for full prioritization.

The model we present for age-prioritization is also relevant to any other ordered risk factor. The variable we use use for *a* could, for example, represent an ordered risk score that combines factors such as age, health status, and risk of exposure. The quantity *𝒟*_*A*_ will be smaller when the concentration of risk is greater. The benefit of taking account of additional risk factors beyond age can be measured by recalculating alternative *𝒟*_*A*_ values for different measures of risk.

Viewed in this way, the quantity 𝒟_*A*_ can be seen as a measure of the concentration of population risk. In fact, graphically it bears a strong resemblance to the Gini index and other measures of concentration, which researchers such as Shkolnikov et al. (2003) have looked at regarding age and mortality. A difference in this case is that it is not just the distribution of deaths that matters, but also the age-distribution of the population.

## Discussion

Strict risk ordering of vaccination is optimal in that it most reduces deaths among those receiving the vaccine (assuming no indirect protection from vaccine roll-out). However, there are other considerations that can and arguably should be taken into account. For example protecting front-line workers, teachers, and others may produce positive benefits for the larger society beyond protecting these individuals. The equity implications of risk-prioritization are also important to consider. More complete analysis accounting for transmission risks and indirect effects (e.g., Bubar et al. (2021)) is also necessary.

Generous eligibility for vaccines is an understandable popular demand, particularly in the face of the slow roll-out and vaccine hesitancy. However, opening eligibility too soon risks losing much of the enormous benefits of prioritization. Even worse, open eligibility in the face of vaccine shortages can result in the least at risk going first, as the same resources that protect individuals from infection and death are used to move to the front of the line. Such “reverse prioritization” would result in even more deaths than the theoretically “neutral” approach.

The politics and logistics of vaccine allocation are complicated. But the demography of the direct effect of vaccination is simple, particularly regarding age: the elderly are at much higher risk but represent only a small fraction of the population. A successful vaccination campaign is not just a question of getting needles into arms. It matters who is vaccinated when. Making every effort to vaccinate the elderly first can save an enormous number of lives.

## Mathematical appendix

### Definitions

*p* The fraction of the population protected from mortality through vaccination, beginning at 0 and ending at 1. A general formulation could have *p*(*t*) take any non-decreasing form. We consider the simple case of a constant rate of vaccination, or *p*(*t*) = *t*, with 0 ≤ *t* ≤ 1.

*d*_0_(*a*) The number of people dying of Covid-19 at each age *a* per unit of time at baseline when no-one is protected. In its normalized form, with 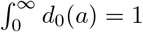,this gives the share of baseline deaths at age *a*.

*F* (*a*) The cumulative distribution function of the population, i.e., the fraction of the population aged less than *a*. 1 − *F* (*a*) is the fraction aged more than *a*.

*A*(*p*) The age above which there is a fraction *p* of the population. Under age-prioritization with constant shares of the population being protected over time, *A*(*p*) is the age above which there is no mortality when a fraction *p* has been been protected.

*𝒟*_*A*_ The ratio of deaths to baseline over the entire course of an age-prioritized vaccination campaign that protects older people first.

*𝒟*_*N*_ The ratio of deaths to baseline over the entire course of an age-neutral vaccination campaign that protects an equal share of each age group each day.

We now calculate the deaths over the full time period of the campaign. Sums of normalized *d*_0_(*a*) (see above) give deaths as a fraction of the baseline total.

Substituting (2) into (3), we have

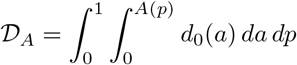

Reversing the order of integration, noting that *p* = 1 − *F* (*a*) is the inverse of *A*(*p*), gives

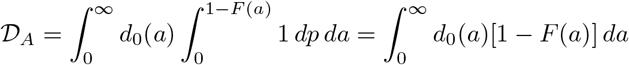

the result in equation (4) of the text.

Similarly, we can consider the deaths-relative-to-baseline over the course of an age-neutral campaign

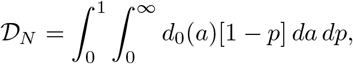

which evaluates to 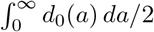, or one-half of the baseline total.

## Data Availability

Publicly available data.
U.S. age distribution from Human Mortality Database (www.mortality.org)
Covid age-specific mortality from Goldstein and Lee (2020)

https://raw.githubusercontent.com/josh-goldstein-git/dempersp_covid_mortality/master/data/cleaned/normalized_nMx_out.csv

## Footnotes

1 This model could be extended to allow for a surface of deaths *d*(*a, t*) that changes with both age and time. Similarly, our assumption that pace of vaccination is constant over the course of the campaign could be relaxed by expressing *p* as a function of time.

## Notes

### Competing Interest Statement

The authors have declared no competing interest.

### Funding Statement

JRG and AM are supported by the Berkeley Population Center (NIH/National Institute of Child Health and Human Development [NICHD] program project P2CHD073964), the Center for the Economics and Demography of Aging (NIH/National Institute on Aging [NIA] grant 5P30AG012839), and Berkeley Formal Demography Workshops (NIH/NICHD grant R25HD08313). J.R.G. is also supported by NIH/NIA grant R01AG058940.

### Author Declarations

No IRB approval required.

